# REACT-2 Round 5: increasing prevalence of SARS-CoV-2 antibodies demonstrate impact of the second wave and of vaccine roll-out in England

**DOI:** 10.1101/2021.02.26.21252512

**Authors:** Helen Ward, Graham Cooke, Matthew Whitaker, Rozlyn Redd, Oliver Eales, Jonathan C Brown, Katharine Collet, Emily Cooper, Anna Daunt, Kathryn Jones, Maya Moshe, Michelle Willicombe, Sophie Day, Christina Atchison, Ara Darzi, Christl A Donnelly, Steven Riley, Deborah Ashby, Wendy S Barclay, Paul Elliott

## Abstract

**Background:** England has experienced high rates of SARS-CoV-2 infection during the COVID-19 pandemic, affecting in particular minority ethnic groups and more deprived communities. A vaccination programme began in England in December 2020, with priority given to administering the first dose to the largest number of older individuals, healthcare and care home workers.

**Methods:** A cross-sectional community survey in England undertaken between 26 January and 8 February 2021 as the fifth round of the REal-time Assessment of Community Transmission-2 (REACT-2) programme. Participants completed questionnaires, including demographic details and clinical and COVID-19 vaccination histories, and self-administered a lateral flow immunoassay (LFIA) test to detect IgG against SARS-CoV-2 spike protein. There were sufficient numbers of participants to analyse antibody positivity after 21 days from vaccination with the PfizerBioNTech but not the AstraZeneca/Oxford vaccine which was introduced slightly later.

**Results:** The survey comprised 172,099 people, with valid IgG antibody results from 155,172. The overall prevalence of antibodies (weighted to be representative of the population of England and adjusted for test sensitivity and specificity) in England was 13.9% (95% CI 13.7, 14.1) overall, 37.9% (37.2, 38.7) in vaccinated and 9.8% (9.6, 10.0) in unvaccinated people. The prevalence of antibodies (weighted) in unvaccinated people was highest in London at 16.9% (16.3, 17.5), and higher in people of Black (22.4%, 20.8, 24.1) and Asian (20.0%, 19.0, 21.0) ethnicity compared to white (8.5%, 8.3, 8.7) people. The uptake of vaccination by age was highest in those aged 80 years or older (93.5%). Vaccine confidence was high with 92.0% (91.9, 92.1) of people saying that they had accepted or intended to accept the offer. Vaccine confidence varied by age and ethnicity, with lower confidence in young people and those of Black ethnicity. Particular concerns were identified around pregnancy, fertility and allergies. In 971 individuals who received two doses of the Pfizer-BioNTech vaccine, the proportion testing positive was high across all age groups. Following a single dose of Pfizer-BioNTech vaccine after 21 days or more, 84.1% (82.2, 85.9) of people under 60 years tested positive (unadjusted) with a decreasing trend with increasing age, but high responses to a single dose in those with confirmed or suspected prior COVID at 90.1% (87.2, 92.4) across all age groups.

**Conclusions:** There is uneven distribution of SARS-CoV-2 antibodies in the population with a higher burden in key workers and some minority ethnic groups, similar to the pattern in the first wave. Confidence in the vaccine programme is high overall although it was lower in some of the higher prevalence groups which suggests the need for improved communication about specific perceived risks. Two doses of Pfizer-BioNTech vaccine, or a single dose following previous infection, confers high levels of antibody positivity across all ages. Further work is needed to understand the relationship between antibody positivity, clinical outcomes such as hospitalisation, and transmission.

## Introduction

England has experienced high rates of SARS-CoV-2 infection during the COVID-19 pandemic, affecting in particular minority ethnic groups and more deprived communities.(1) The REal-time Assessment of Community Transmission-2 (REACT-2) study is a community survey to measure the prevalence of antibodies to the SARS-CoV-2 virus among adults in England. We have previously reported the extent and variation in antibody prevalence after the first wave of COVID-19 including the unequal risk by region, occupation and ethnicity.(1) The overwhelming majority of cases in the first wave occurred in a relatively short period in March and April 2020(1), leading to the first national lockdown, after which we reported waning of antibody prevalence in the population from 6.0% in June to 4.4% in September 2020.(2)

The emergence of the second UK wave of infections was first evident in September 2020,(3) and continued with widespread community transmission despite a three-week second lockdown in England in November 2020.(4) This was followed by a rapid rise in infections associated with the new and more transmissible B1.1.17 variant,(5) which led to a third national lockdown in England in January 2021.

The UK SARS-CoV-2 vaccination programme delivered its first dose in December 2020. Prioritisation followed recommendations from the Joint Committee on Vaccination and Immunization (JCVI), closely aligned to the WHO Roadmap.(6) Implementation of the programme has been rapid, with 15 million people in England receiving at least one dose of vaccine by the third week of February.(7) There are some early signs that the programme, initially targeting people over-70 years of age, health and care workers, older adults, care home residents and clinically extremely vulnerable people, is having an impact on hospitalisations, mortality and, possibly, transmission. (8,9)

Here we report the results of REACT-2 round 5 carried out between 26 January and 8 February 2021, which includes some people who have received one or more doses of a COVID-19 vaccine. We report the overall prevalence of positivity for SARS-CoV-2 IgG antibodies in the community in vaccinated and unvaccinated individuals, the impact of vaccination on antibody status, and confidence in vaccination across the population.

## Methods

REACT-2 is a repeated cross-sectional community survey of adults in England. The protocol and earlier results have been published.(1,2,10) In brief, each round of study includes a random, non-overlapping community sample from the adult population 18 years and older, using a self-administered lateral flow immunoassay test (LFIA) at home. Tests were sent to named individuals randomly selected from the National Health Service (NHS) patient list that includes anyone registered with a General Practitioner in England and covers almost the entire population. Personalized invitations were sent to 600,000 individuals aged 18 years and above to achieve similar numbers of respondents in each of 315 lower-tier local authority (LTLA) areas. Participants registered via an online portal or by telephone with registration closed after ∼200,000 people had signed up. Response rates are provided in Supplementary Data (Table S1).

Survey instruments are available on the study website^1^. Those who registered were sent a test kit, including a self-administered point-of-care LFIA test and instructions by post, with link to an on-line instruction video. Participants completed a short registration questionnaire (online/telephone) and a further survey upon completion of their self-test, including information on demographics, household composition, symptoms, and history of COVID-19; participants also uploaded a photograph of the result. In round 5, questions were additionally included on vaccination, covering whether or not participants had received a vaccine, and if so the date and number of doses. Those unvaccinated were asked whether they had been invited and their response. For those who had not yet been offered a vaccine we asked about their intention to accept. People who reported being unsure or who would decline vaccination were asked to select from a list of possible reasons for hesitancy based on issues identified from previous research,(11) with the additional option of free-text responses.

The LFIA (Fortress Diagnostics, Northern Ireland) targeting the spike protein was selected following evaluation of performance characteristics (sensitivity and specificity) against pre-defined criteria for detection of IgG,(12) and extensive public involvement and user testing.(13) The LFIA has a clinical sensitivity on finger-prick blood (self-read) for IgG antibodies following natural infection estimated at 84.4% (70.5, 93.5) in RT-PCR confirmed cases in healthcare workers, and specificity 98.6% (97.1, 99.4) in pre-pandemic sera.(12,14) The LFIA detects immune responses to the spike (S) protein targeted by available vaccines. The performance of the LFIA was evaluated using the sera of vaccinated healthcare workers recruited between 23 December 2020 and 31 January 2021. Seventy-two individuals were sampled before receiving a single 30μl dose of Pfizer-BioNTech (BNT162b2) vaccine and again 21-25 days following vaccination. Participants were tested for antibodies to SARS-CoV-2 spike (anti-S) protein using the Abbott IgG Quant II chemiluminescent immunoassay (threshold value for positivity 50 AU/ml). Thirty-one individuals from this set were assessed for neutralisation using live SARS-CoV-2 virus (SARS-CoV-2/England/IC19/2020) neutralisation assays on Vero-E6 cells as described previously.(2)

### Data analyses

Data were analysed using the statistical package R version 4.0.0.(15)

Prevalence was calculated as the proportion of individuals with a positive IgG result on the LFIA. For analyses at population level (but not for individual vaccine response) we adjusted for test performance using:

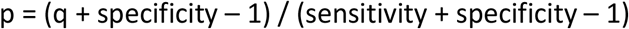

where p = adjusted proportion positive, q = observed proportion positive.(16) Prevalence estimates at national level were weighted for age, sex, region, ethnicity and deprivation using Index of Multiple Deprivation (IMD) quintiles(17) to account for the geographic sample design and for variation in response rates, so as to be representative of the population (18+ years) of England.

We report “vaccine confidence”, defined as accepting or planning to accept the vaccine offer, and analyse reasons for vaccine hesitancy including a thematic analysis of free-text responses.

We obtained research ethics approval from the South Central-Berkshire B Research Ethics Committee (IRAS ID: 283787), and MHRA approval for use of the LFIA for research purposes only. The REACT Public Advisory Panel provides regular review of the study processes and results. The healthcare worker study was approved by the Health Research Authority, Research Ethics Committee (Reference: 20/WA/0123).

## Results

Round 5 includes questionnaire responses from 172,099 people with valid IgG antibody results from 155,172.

### Prevalence

The overall prevalence of antibodies (weighted, adjusted) in England was 13.9% (95% CI 13.7, 14.1) overall, 37.9% (37.1, 38.7) in vaccinated and 9.8% (9.6, 10.0) in unvaccinated people (increased from 5.6% (5.4, 5.7) in Round 4 in November 2020, Table 1). The prevalence in vaccinated people reflects the recent roll-out of the programme with many people having received their first dose in the preceding three weeks and who would not have had time to produce detectable antibodies. The prevalence of antibodies (weighted) in unvaccinated people was highest in London at 16.9% (16.3, 17.5), and higher in people of Black (22.4%, 20.8, 24.1) and Asian (20.0%, 19.0, 21.0) compared to white (8.5%, 8.3, 8.7) ethnicity. Prevalence was highest in those aged 18-29 years at 14.5% (14.0, 15.0), higher in females (10.5%, 10.2, 10.8) than males (9.1%, 8.8, 9.3), and higher in people living in the most deprived IMD quintile at 12.3% (11.8, 12.8) compared to 7.7% (7.3, 8.1) in the least. (Table 2) An epidemic curve constructed from date of onset of symptoms in unvaccinated people who were IgG positive shows that the second wave grew more slowly in September to November than the first wave in March-April, and then accelerated in December 2020. (Figure 1)

**Table 1:**
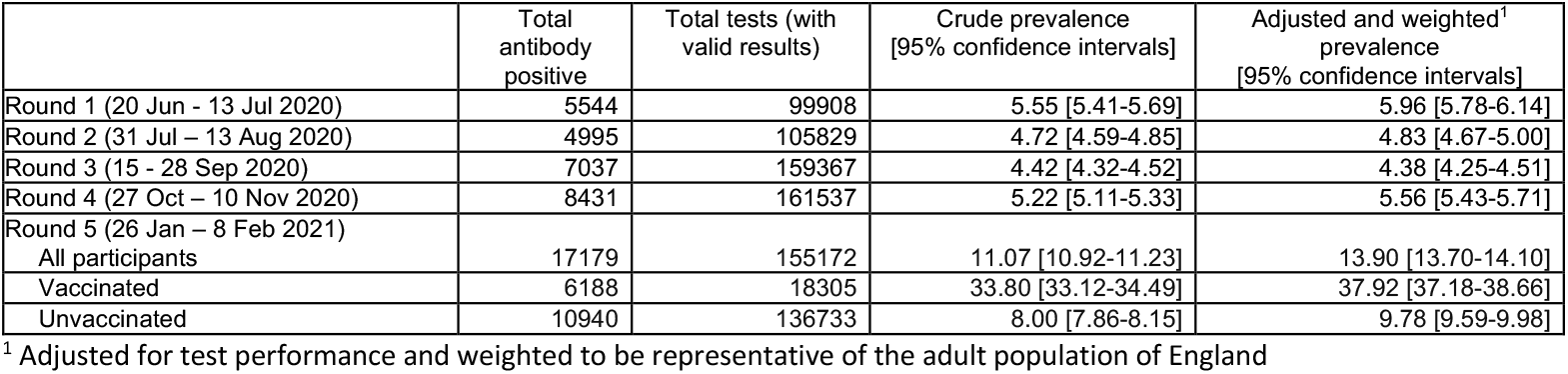
REACT-2: Community prevalence of IgG antibodies to SARS-CoV-2 in adults in England, adjusted and weighted, June 2020 – Feb 2021.

|  | Total antibody positive | Total tests (with valid results) | Crude prevalence<br>[95% confidence intervals] | Adjusted and weighted<sup>1</sup> prevalence<br>[95% confidence intervals] |
| --- | --- | --- | --- | --- |
| Round 1 (20 Jun - 13 Jul 2020) | 5544 | 99908 | 5.55 [5.41-5.69] | 5.96 [5.78-6.14] |
| Round 2 (31 Jul – 13 Aug 2020) | 4995 | 105829 | 4.72 [4.59-4.85] | 4.83 [4.67-5.00] |
| Round 3 (15 - 28 Sep 2020) | 7037 | 159367 | 4.42 [4.32-4.52] | 4.38 [4.25-4.51] |
| Round 4 (27 Oct – 10 Nov 2020) | 8431 | 161537 | 5.22 [5.11-5.33] | 5.56 [5.43-5.71] |
| Round 5 (26 Jan – 8 Feb 2021) |  |  |  |  |
| All participants | 17179 | 155172 | 11.07 [10.92-11.23] | 13.90 [13.70-14.10] |
| Vaccinated | 6188 | 18305 | 33.80 [33.12-34.49] | 37.92 [37.18-38.66] |
| Unvaccinated | 10940 | 136733 | 8.00 [7.86-8.15] | 9.78 [9.59-9.98] |

<sup>1</sup> Adjusted for test performance and weighted to be representative of the adult population of England

**Table 2:** IgG prevalence, adjusted and weighted, in unvaccinated* people by sex, age, region, ethnicity and deprivation.

| Question | Category | Total antibody positive | Total tests (with valid results) | Crude prevalence | Prevalence adjusted for test | Weighted prevalence |
| --- | --- | --- | --- | --- | --- | --- |
|  | All respondents | 10965 | 137095 | 8 (7.9-8.1) | 7.9 (7.8-8.1) | 9.8 (9.6-10) |
| Sex | Female | 6381 | 75106 | 8.5 (8.3-8.7) | 8.5 (8.3-8.8) | 10.5 (10.2-10.8) |
| Sex | Male | 4583 | 61985 | 7.4 (7.2-7.6) | 7.2 (7-7.5) | 9.1 (8.8-9.3) |
| Age group | 18-29 | 1985 | 16790 | 11.8 (11.3-12.3) | 12.6 (12-13.2) | 14.5 (14-15) |
| Age group | 30-39 | 1932 | 22427 | 8.6 (8.3-9) | 8.7 (8.3-9.1) | 9.9 (9.5-10.4) |
| Age group | 40-49 | 2213 | 26486 | 8.4 (8-8.7) | 8.4 (8-8.8) | 9.6 (9.2-10.1) |
| Age group | 50-59 | 2528 | 31341 | 8.1 (7.8-8.4) | 8 (7.7-8.4) | 9.1 (8.7-9.6) |
| Age group | 60-69 | 1740 | 27875 | 6.2 (6-6.5) | 5.8 (5.5-6.2) | 6.9 (6.5-7.3) |
| Age group | 70-79 | 547 | 11957 | 4.6 (4.2-5) | 3.8 (3.4-4.3) | 4.5 (4.1-5.1) |
| Age group | 80+ | 20 | 219 | 9.1 (6-13.7) | 9.3 (5.5-14.8) | 10.3 (7.4-14.1) |
| Ethnicity | Asian | 577 | 4005 | 14.4 (13.4-15.5) | 15.7 (14.4-17) | 20.0 (19-21) |
| Ethnicity | Black | 171 | 987 | 17.3 (15.1-19.8) | 19.2 (16.5-22.2) | 22.4 (20.8-24.1) |
| Ethnicity | Mixed | 176 | 1734 | 10.1 (8.8-11.7) | 10.5 (8.9-12.4) | 12.6 (10.8-14.6) |
| Ethnicity | Other | 136 | 984 | 13.8 (11.8-16.1) | 15 (12.5-17.7) | 17.6 (15.3-20.2) |
| Ethnicity | White | 9819 | 128454 | 7.6 (7.5-7.8) | 7.5 (7.3-7.7) | 8.5 (8.3-8.7) |
| IMD quintile | 1 - least deprived | 1330 | 13250 | 10 (9.5-10.6) | 10.4 (9.8-11) | 12.3 (11.8-12.8) |
|  | 2 | 2003 | 21769 | 9.2 (8.8-9.6) | 9.4 (8.9-9.9) | 11.6 (11.1-12) |
|  | 3 | 2258 | 29560 | 7.6 (7.3-7.9) | 7.5 (7.2-7.9) | 8.8 (8.4-9.2) |
|  | 4 | 2602 | 34012 | 7.7 (7.4-7.9) | 7.5 (7.2-7.9) | 8.5 (8.1-8.9) |
|  | 5 - least deprived | 2772 | 38504 | 7.2 (6.9-7.5) | 7 (6.7-7.3) | 7.7 (7.3-8.1) |
| Region | East Midlands | 1271 | 17914 | 7.1 (6.7-7.5) | 6.9 (6.4-7.3) | 7.8 (7.3-8.5) |
|  | East of England | 1528 | 19567 | 7.8 (7.4-8.2) | 7.7 (7.3-8.2) | 8.3 (7.8-8.9) |
|  | London | 1676 | 12284 | 13.6 (13-14.3) | 14.8 (14-15.5) | 16.9 (16.3-17.5) |
|  | North East | 441 | 5050 | 8.7 (8-9.5) | 8.8 (7.9-9.8) | 9 (8.2-9.8) |
|  | North West | 1568 | 15992 | 9.8 (9.4-10.3) | 10.1 (9.6-10.7) | 11.7 (11.2-12.3) |
|  | South East | 2095 | 30279 | 6.9 (6.6-7.2) | 6.6 (6.3-7) | 7.4 (7-7.8) |
|  | South West | 675 | 13870 | 4.9 (4.5-5.2) | 4.2 (3.8-4.6) | 4.8 (4.4-5.3) |
|  | West Midlands | 1031 | 13058 | 7.9 (7.4-8.4) | 7.8 (7.3-8.4) | 10.3 (9.7-10.9) |
|  | Yorkshire and The Humber | 680 | 9081 | 7.5 (7-8) | 7.3 (6.7-8) | 8.8 (8.2-9.4) |

\*487 respondents reported a date of vaccination after their swab test date but before they completed the survey. These respondents are classified as unvaccinated in all prevalence calculations.

**Figure 1:**
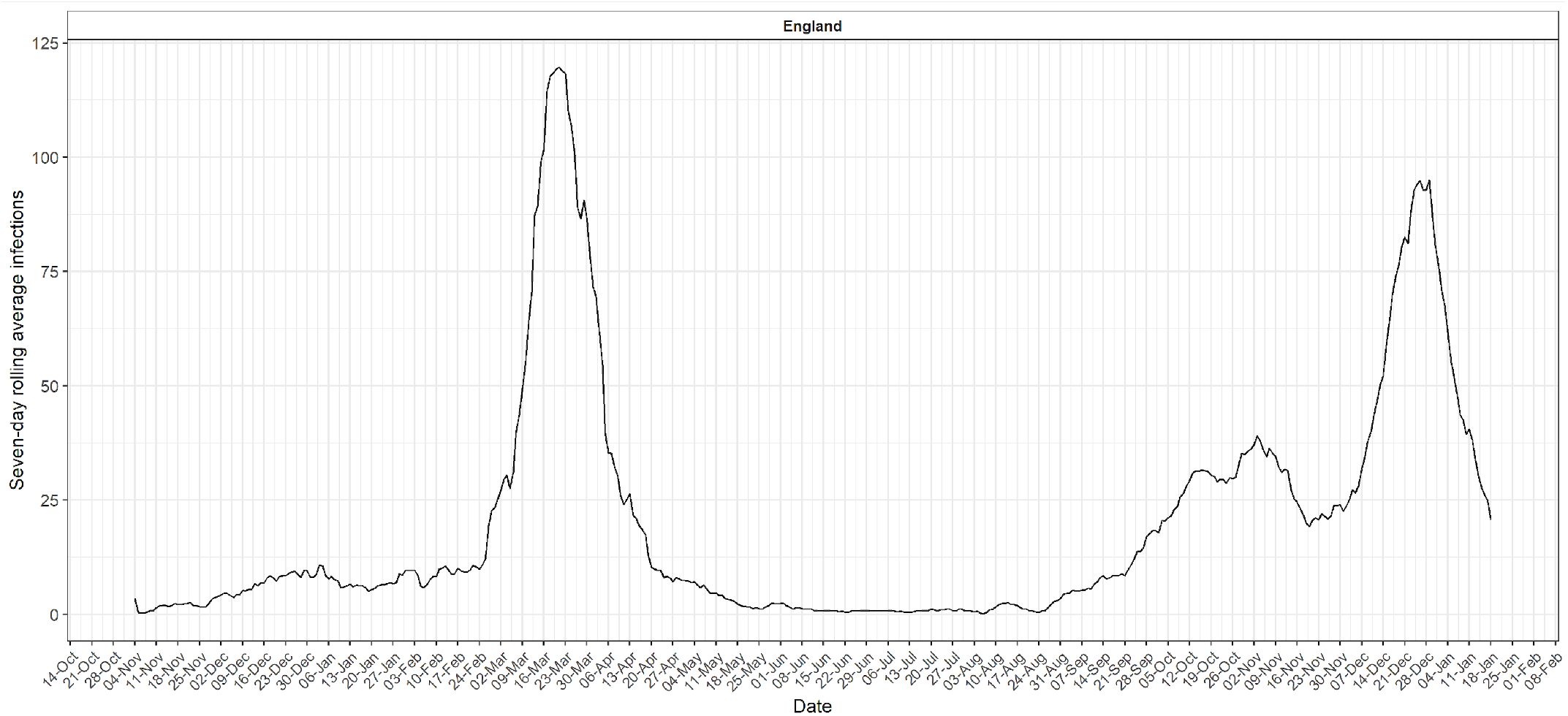
Reconstructed epidemic curve from number of symptomatic infections per week, by date of onset in antibody positive participants reporting symptoms.

Prevalence of antibodies by employment type (unweighted) for unvaccinated participants was highest in healthcare and care home workers at 21.9% (20.0, 23.9) and 24.2% (19.8, 29.1) respectively. The prevalence among those working in public transport was 12.2%, (10.1, 14.7), police and prison 11.9% (9.9, 14.1), education 11.4% (10.7, 12.2), childcare 11.4% (9.0, 14.3), and personal care 11.1% (9.0, 13.4), higher than in non key-workers (7.8%, 7.6, 8.1). Other unvaccinated groups with high antibody prevalence included those living in larger households of 7 or more people at 18.4% (15.8, 21.3) and people of Bangladeshi (25.4%, 19.1, 32.8), African (23.4%, 19.6, 27.8), and Pakistani (21.9%, 18.1, 26.1) ethnic sub-categories. (Table 3)

**Table 3:**
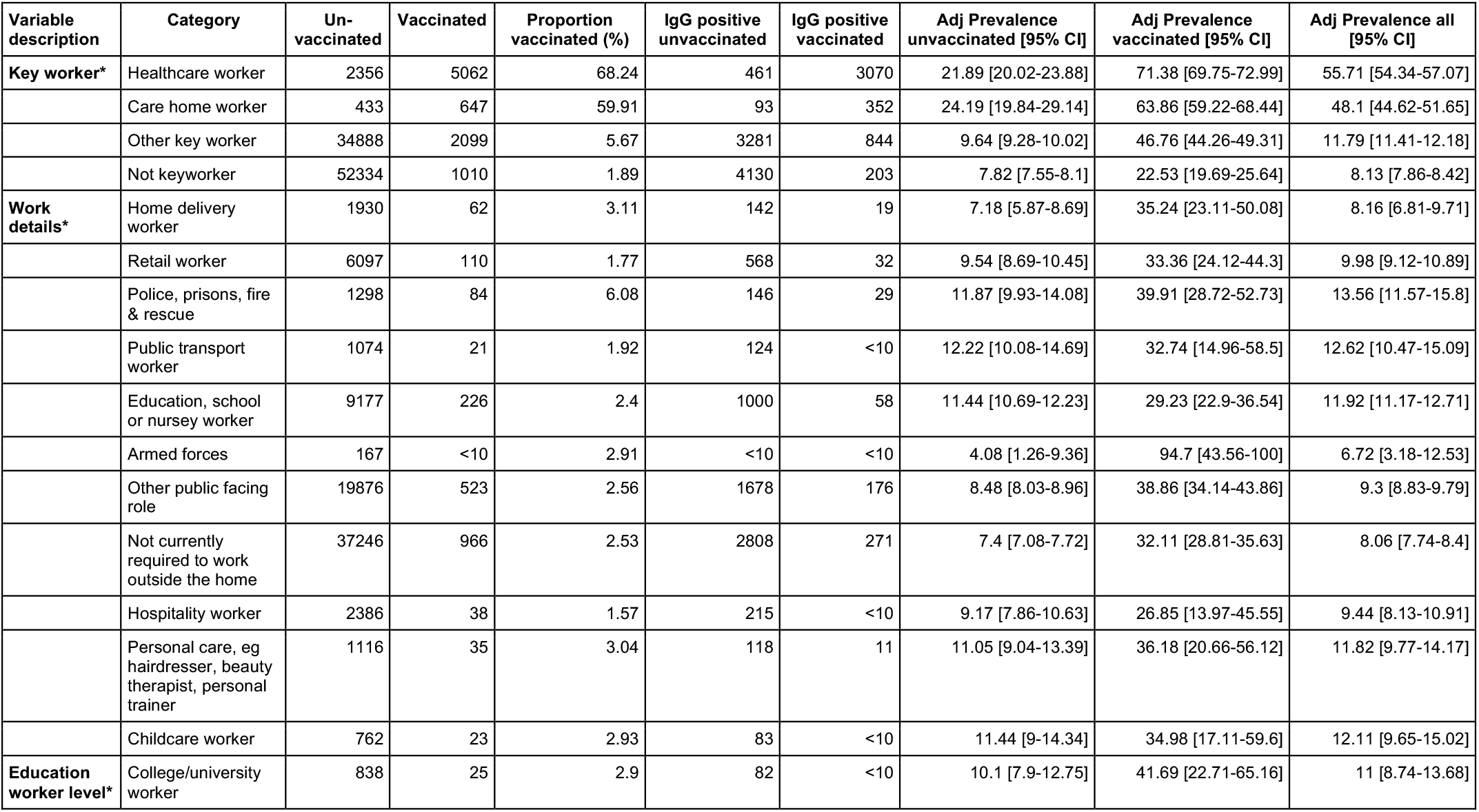
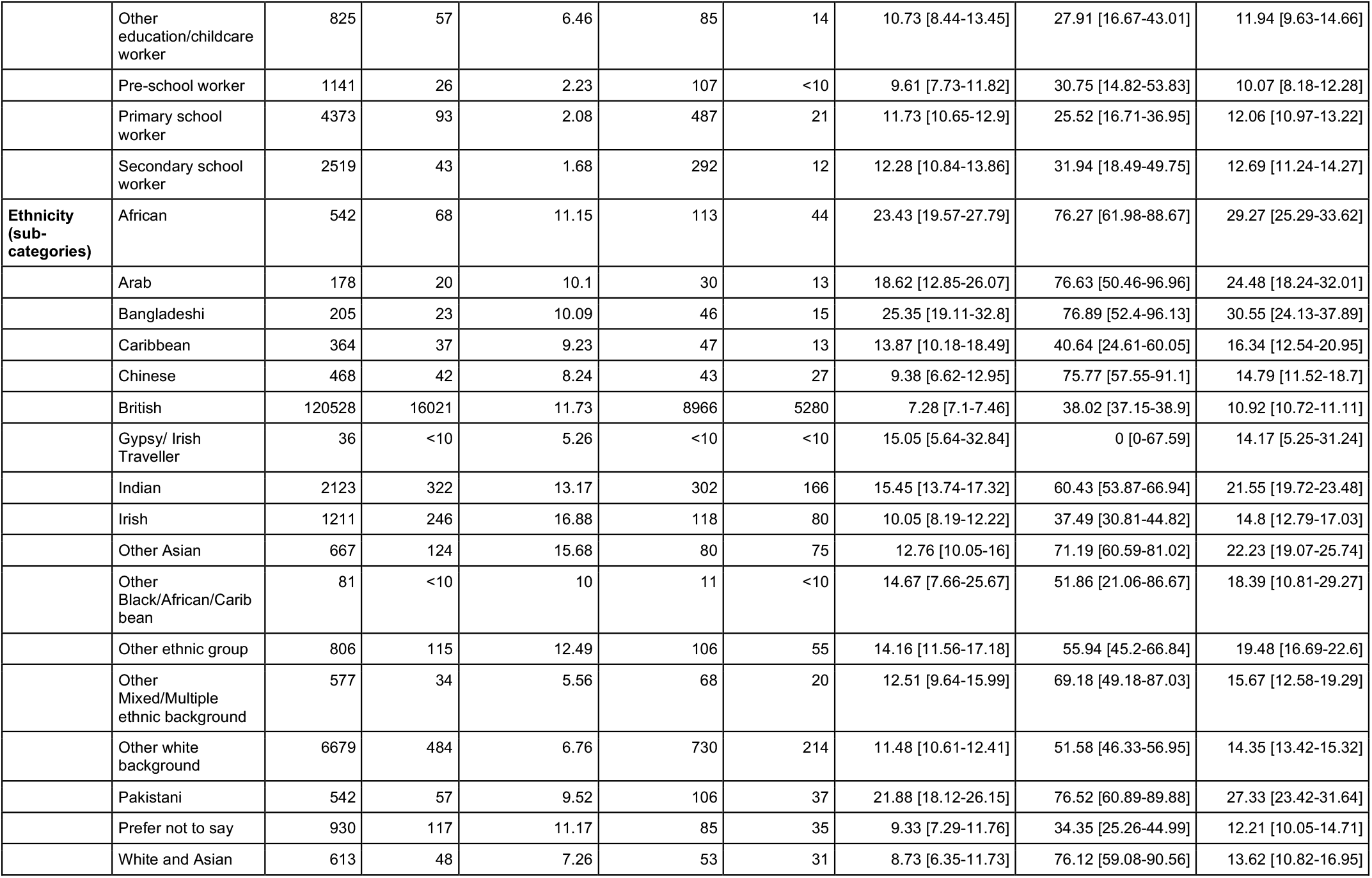
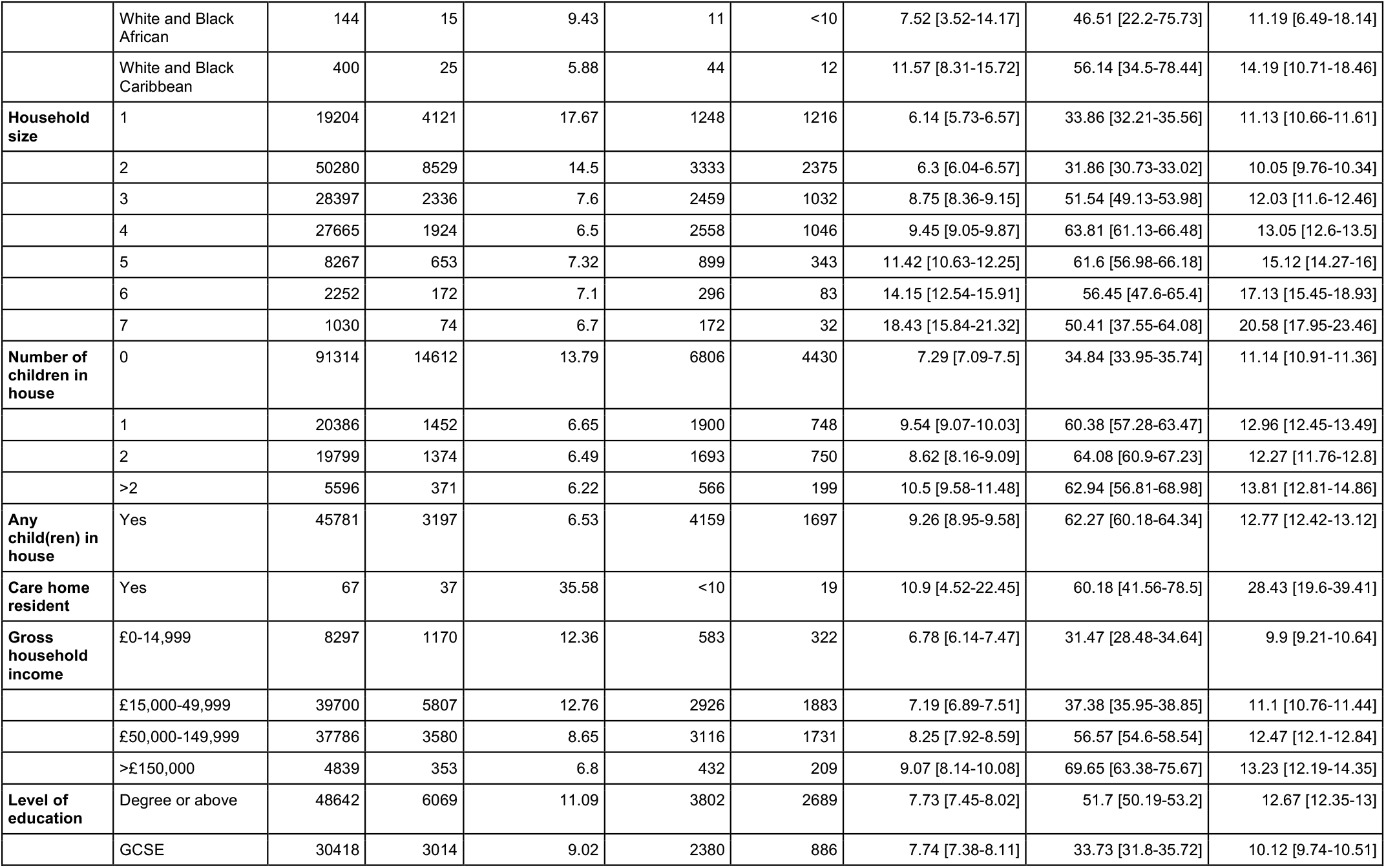
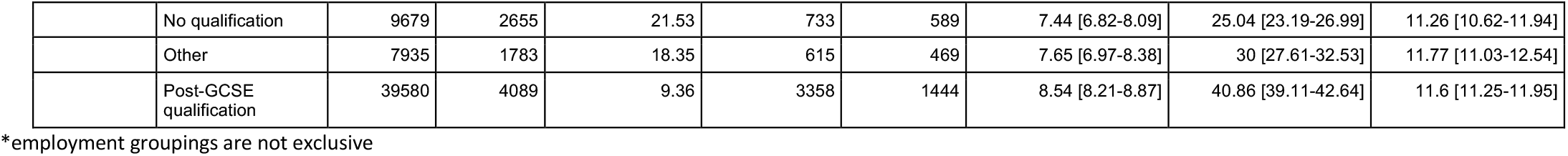
Vaccine coverage, and IgG prevalence, adjusted and unweighted, by vaccination status by occupational detail, ethnic sub-groups, household size and composition, income and education.

### Antibody response after vaccination

Overall, 18,086 individuals with valid antibody results reported prior vaccination with either Pfizer-BioNTech or AstraZeneca/Oxford (ChAdOX1) nCoV-19 vaccines.(18) At the time of this study 5,266 reported vaccination with AstraZeneca/Oxford, but only five had received two doses and the majority (5,084/5,266, 95.6%) had only received a single dose within the last 21 days. Accordingly, further analysis was limited to those receiving the Pfizer-BioNTech (with 12,820 receiving at least one dose).

The uptake of vaccination by age was highest in those aged 80 years or older (93.4%), followed by those aged 70-79 years (33.7%), and by occupation highest in healthcare workers (68.2%) and care home workers (59.9%). (Table 3) In 971 individuals who received two doses of the Pfizer-BioNTech vaccine, the proportion testing positive was high across all age groups, at 91.1% (88.5, 97.1) overall. Following a single dose of Pfizer-BioNTech vaccine after 21 days or more, 84.1% (82.2, 85.9) of people under 60 years tested positive (unadjusted) with a decreasing trend with increasing age, but high responses to a single dose in those with confirmed or suspected prior COVID at 88.8% (85.9, 91.2) overall. (Table 4, Figure 2) The apparent higher positivity in people with prior COVID-19 was present in all age groups. (Table 5, Figure 3) Antibody positivity appeared to plateau after four to five weeks in all age groups following a single Pfizer-BioNTech vaccine. (Table 5, Figure3).

**Table 4:** IgG positivity 21 days or more after one and two Pfizer/BioNTech doses, by age group.

|  | Pfizer single dose, >21 days earlier |  |  |  |  | Pfizer two doses |  |  |  |  |
| --- | --- | --- | --- | --- | --- | --- | --- | --- | --- | --- |
| Category | Positive | Total | Prevalence | Lower CI | Upper | Positive | Total | Prevalence | Lower | Upper |
| 18-29 | 213 | 225 | 94.7 | 90.9 | 96.9 | 30 | 30 | 100.0 | 91.7 | 100.0 |
| 30-39 | 270 | 300 | 90.0 | 86.1 | 92.9 | 48 | 48 | 100.0 | 94.7 | 100.0 |
| 40-49 | 358 | 425 | 84.2 | 80.5 | 87.4 | 104 | 108 | 96.3 | 90.9 | 98.6 |
| 50-59 | 462 | 599 | 77.1 | 73.6 | 80.3 | 118 | 128 | 92.2 | 86.2 | 95.7 |
| 60-69 | 221 | 313 | 70.6 | 65.3 | 75.4 | 70 | 73 | 95.9 | 88.6 | 98.6 |
| 70-79 | 148 | 304 | 48.7 | 43.1 | 54.3 | 38 | 41 | 92.7 | 80.6 | 97.5 |
| 80+ | 293 | 845 | 34.7 | 31.5 | 37.9 | 477 | 543 | 87.8 | 84.8 | 90.3 |

**Table 5:** IgG positivity by days after single dose of Pfizer/BioNTech vaccine, by age, sex and clinical history.

|  | IgG positivity 5 (95% confidence interval) |  |  |  |  |
| --- | --- | --- | --- | --- | --- |
| Category | <21 days | 21-27 days | 28-34 days | 35-41 days | >=42 days |
| All respondents | 30 (29.1-31) | 68.2 (65.8-70.6) | 67.7 (64-71.3) | 59.9 (56.2-63.5) | 56.7 (50.6-62.6) |
| 18-29 | 57.1 (53-61.1) | 92.5 (86.4-96) | 100 (94.4-100) | 95.1 (83.9-98.7) | 94.4 (74.2-99) |
| 30-39 | 51 (47.6-54.4) | 91.6 (86.1-95) | 94.2 (86-97.7) | 85.4 (72.8-92.8) | 79.3 (61.6-90.2) |
| 40-49 | 44.5 (41.5-47.6) | 84.4 (78.9-88.6) | 85.6 (76.8-91.4) | 84.8 (76.1-90.7) | 78.1 (61.2-89) |
| 50-59 | 40.3 (37.7-42.9) | 83.1 (78.5-86.8) | 79.7 (71.5-85.9) | 66.4 (57.4-74.3) | 62.1 (49.2-73.4) |
| 60-69 | 28 (25.2-31) | 70.6 (63.1-77) | 69 (57.5-78.6) | 76.3 (64-85.3) | 60 (38.7-78.1) |
| 70-79 | 14 (12.8-15.3) | 50.5 (43.7-57.2) | 42.1 (30.2-55) | 50 (32.6-67.4) | 45.5 (21.3-72) |
| 80+ | 20.8 (18.6-23.2) | 30.3 (25.3-35.8) | 38 (30.9-45.5) | 38 (32.6-43.7) | 32.3 (23.6-42.3) |
| Female | 34.6 (33.4-35.9) | 73.9 (71.1-76.5) | 72.4 (68.1-76.3) | 66.2 (61.7-70.4) | 64.5 (56.9-71.3) |
| Male | 21.6 (20.2-23.1) | 56 (51.4-60.4) | 55.4 (47.8-62.7) | 47.3 (40.9-53.8) | 43.2 (33.7-53.2) |
| Not clinically vulnerable | 31.2 (30.2-32.2) | 70.1 (67.6-72.5) | 70.6 (66.7-74.3) | 61.3 (57.3-65.1) | 58.8 (52.3-65.1) |
| Clinically vulnerable/ advised to shield | 20.2 (17.7-22.9) | 48.4 (39.8-57.1) | 41.9 (30.5-54.3) | 50.6 (40.3-60.8) | 42.9 (28-59.1) |
| COVID suspected or confirmed | 62.1 (59.4-64.8) | 90.1 (85.8-93.2) | 93.1 (87-96.5) | 86 (77.9-91.5) | 91.9 (78.7-97.2) |
| No COVID | 24.8 (23.8-25.8) | 63.6 (60.9-66.3) | 61.9 (57.5-66) | 55.4 (51.3-59.4) | 50.9 (44.4-57.4) |

**Figure 2.**
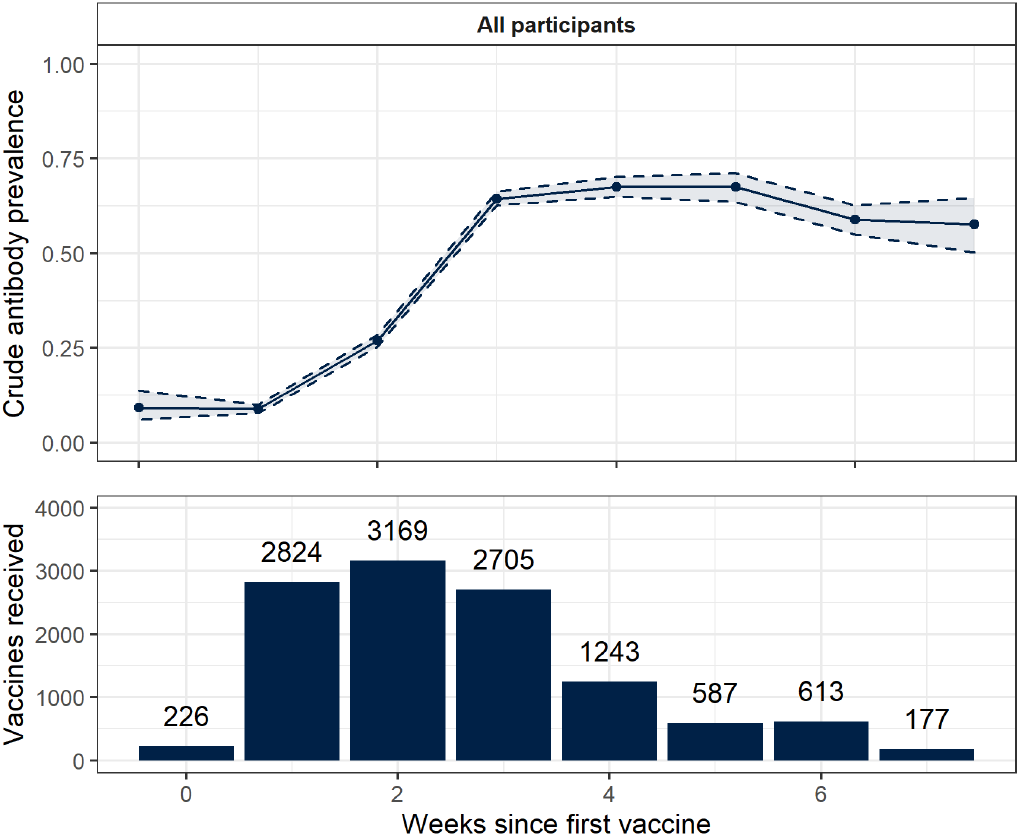
Unadjusted antibody positivity up to 7 weeks following single Pfizer-BioNTech vaccination (aggregated by week) Lower panel shows counts of vaccines received, aggregated by number of weeks since the vaccine was received. Upper plot shows unadjusted proportions of respondents who tested positive for antibodies, aggregated by number of weeks since the vaccine was received. Binomial confidence intervals constructed using the Wilson method are shown.

**Figure 3.**
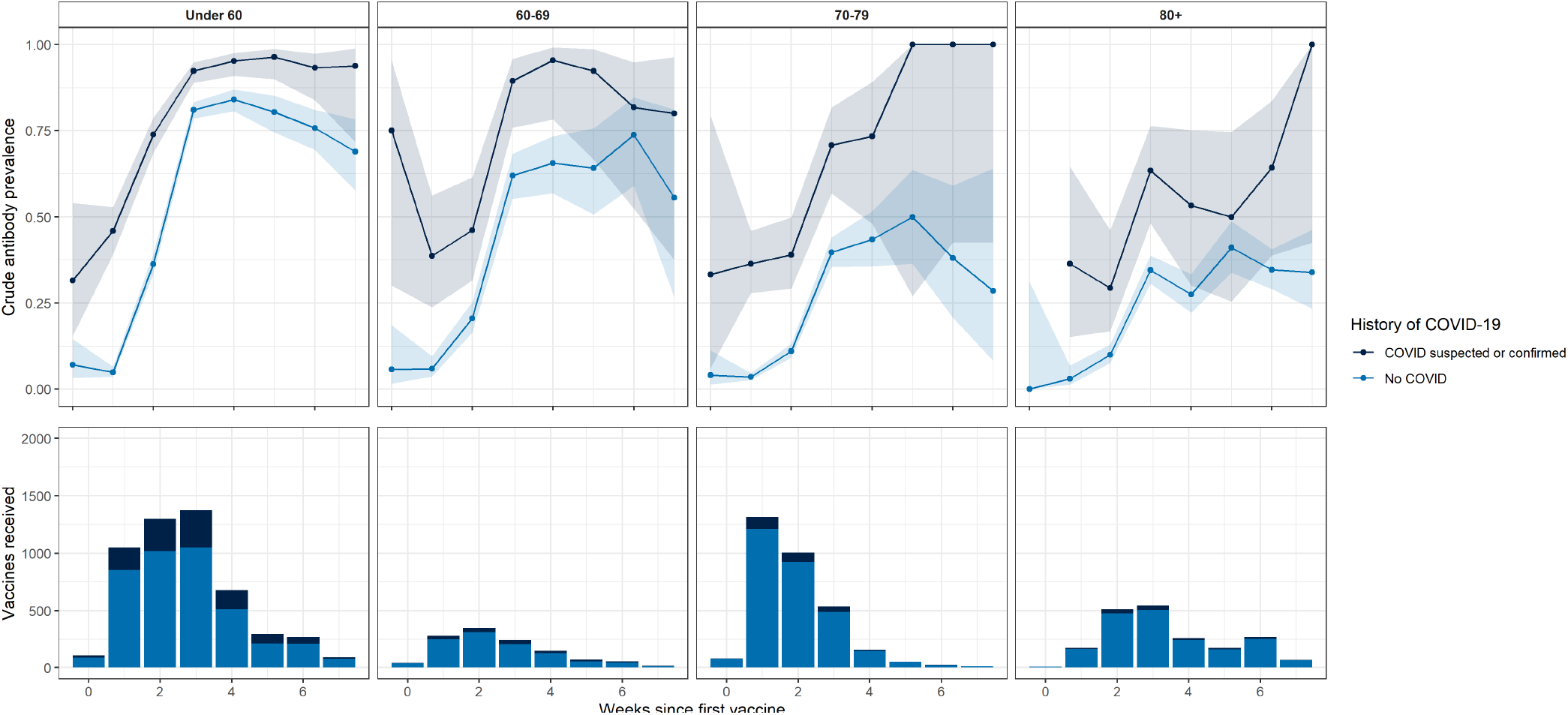
IgG positivity with time since single dose of Pfizer-BioNTech vaccine by age and prior COVID-19 status. Lower panels show counts of vaccines received, aggregated by number of weeks since the vaccine was received. Upper plots show unadjusted proportions of respondents who tested positive for antibodies, aggregated by number of weeks since the vaccine was received, separately for those with no history of COVID-19 and those with confirmed or suspected COVID-19. Binomial confidence intervals constructed using the Wilson method are shown.

In the sera from the cohort of healthcare workers, a comparison with the Abbott anti-S ELISA found the LFIA was positive for IgG in 28/30 samples with antibody levels > 50 U/ml and 29/30 sera with positive LFIA results showed evidence of live virus neutralisation. (Figure 5)

**Figure 5.**
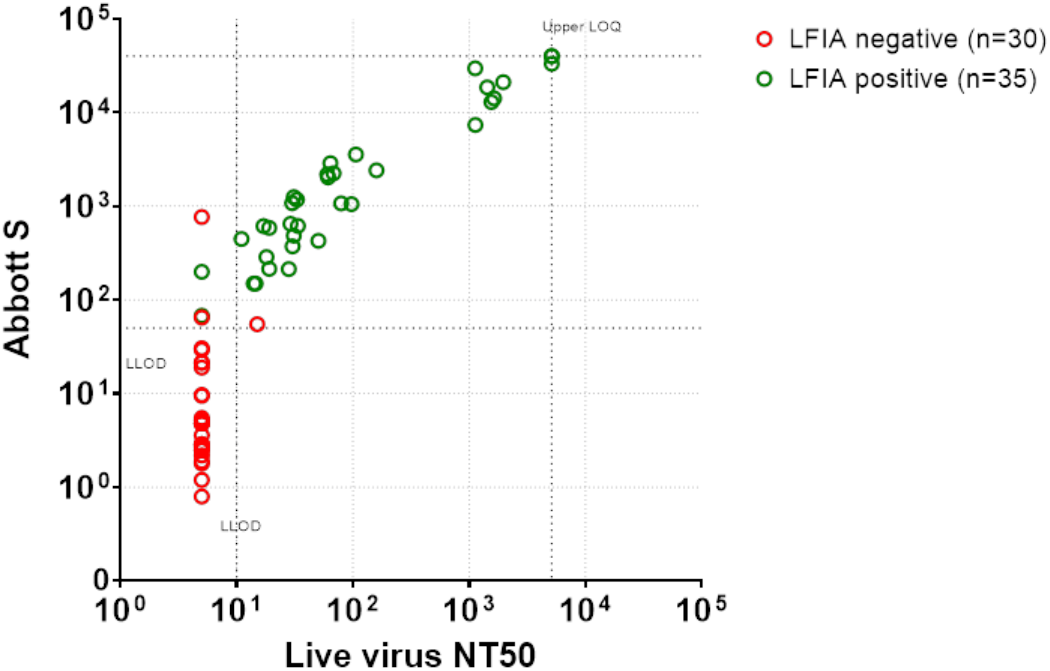
Association between LFIA (Fortress) live virus neutralisation and anti-S IgG levels. Association in vaccinated healthcare workers between LFIA (Fortress), live virus neutralisation and anti-S IgG levels as measured by Abbott Quant II chemiluminescent immunoassays. All samples had lateral flow assays that were either positive (red circles) or negative (green circles). See supplementary material for methods.

### Vaccine confidence

Vaccine confidence was high as shown in Table 6, with 92.0% (91.9, 92.1) of people saying that they had accepted or intended to accept the offer. This varied by age, being higher in older groups with 99.0% (98.6, 99.2) of those aged 80 years or older compared to 83.4%, (82.8,83.9) of 18 to 29 year olds. Confidence was lower in females 90.7% (90.5, 90.9) than males 93.6% (93.4, 93.8). Vaccine confidence also varied by ethnicity, being highest in those of white ethnicity at 92.6% (92.5, 92.7) and lowest in those of Black ethnicity at 72.5% (70.1, 74.8). Vaccine confidence was slightly lower in care home workers at 88.5% (86.7, 90.2) than healthcare workers at 92.1% (91.5, 92.7). (Figure 4) Among those reporting that they would decline or were unsure about vaccine, the most commonly reported reasons for decline/ hesitancy – based on responses to a predefined list where multiple response were possible – were wanting to wait and see how the vaccine works, worries about long-term health effects and about side effects. Additional free text comments showed common concerns around current and planned pregnancy, future fertility and specific allergies or comorbidities.

**Table 6:** REACT-2: Vaccine confidence among adults in England based on actual and intended response to invitation.

| Variable | Category | accepted / would accept vaccine | Declined / would decline vaccine | Don't know / prefer not to say |
| --- | --- | --- | --- | --- |
| **All participants** |  | 158315 (92%, [91.9-92.1]) | 2476 (1.4%, [1.4-1.5]) | 11308 (6.6%, [6.5-6.7]) |
| **Sex** | Female | 86616 (90.7%, [90.5-90.9]) | 1582 (1.7%, [1.6-1.7]) | 7288 (7.6%, [7.5-7.8]) |
|  | Male | 71696 (93.6%, [93.4-93.8]) | 894 (1.2%, [1.1-1.2]) | 4019 (5.2%, [5.1-5.4]) |
| **Age group** | 18-29 | 16170 (83.4%, [82.8-83.9]) | 655 (3.4%, [3.1-3.6]) | 2574 (13.3%, [12.8-13.8]) |
|  | 30-39 | 22409 (84.9%, [84.5-85.4]) | 768 (2.9%, [2.7-3.1]) | 3205 (12.1%, [11.8-12.5]) |
|  | 40-49 | 28461 (90%, [89.7-90.3]) | 470 (1.5%, [1.4-1.6]) | 2687 (8.5%, [8.2-8.8]) |
|  | 50-59 | 35455 (94.3%, [94.1-94.6]) | 338 (0.9%, [0.8-1]) | 1796 (4.8%, [4.6-5]) |
|  | 60-69 | 31958 (96.9%, [96.7-97.1]) | 165 (0.5%, [0.4-0.6]) | 848 (2.6%, [2.4-2.7]) |
|  | 70-79 | 20079 (98.8%, [98.7-99]) | 63 (0.3%, [0.2-0.4]) | 175 (0.9%, [0.7-1]) |
|  | 80 | 3783 (99%, [98.6-99.2]) | 17 (0.4%, [0.3-0.7]) | 23 (0.6%, [0.4-0.9]) |
| **Ethnicity** | Asian | 4927 (87.6%, [86.7-88.4]) | 98 (1.7%, [1.4-2.1]) | 601 (10.7%, [9.9-11.5]) |
|  | Black | 973 (72.5%, [70.1-74.8]) | 68 (5.1%, [4-6.4]) | 301 (22.4%, [20.3-24.7]) |
|  | Mixed | 1716 (83.1%, [81.4-84.6]) | 62 (3%, [2.3-3.8]) | 288 (13.9%, [12.5-15.5]) |
|  | Other | 1217 (84.4%, [82.4-86.2]) | 37 (2.6%, [1.9-3.5]) | 188 (13%, [11.4-14.9]) |
|  | White | 148485 (92.6%, [92.5-92.7]) | 2164 (1.3%, [1.3-1.4]) | 9720 (6.1%, [5.9-6.2]) |
| **Region** | East Midlands | 20519 (92.5%, [92.2-92.9]) | 337 (1.5%, [1.4-1.7]) | 1318 (5.9%, [5.6-6.3]) |
|  | East of England | 22730 (92.1%, [91.7-92.4]) | 353 (1.4%, [1.3-1.6]) | 1607 (6.5%, [6.2-6.8]) |
|  | London | 14050 (87.8%, [87.3-88.3]) | 338 (2.1%, [1.9-2.3]) | 1620 (10.1%, [9.7-10.6]) |
|  | North East | 5847 (91.7%, [91-92.3]) | 89 (1.4%, [1.1-1.7]) | 441 (6.9%, [6.3-7.6]) |
|  | North West | 18618 (91.5%, [91.2-91.9]) | 319 (1.6%, [1.4-1.7]) | 1401 (6.9%, [6.5-7.2]) |
|  | South East | 34847 (92.7%, [92.4-92.9]) | 457 (1.2%, [1.1-1.3]) | 2303 (6.1%, [5.9-6.4]) |
|  | South West | 16007 (93.4%, [93-93.8]) | 188 (1.1%, [1-1.3]) | 942 (5.5%, [5.2-5.8]) |
|  | West Midlands | 15150 (92.3%, [91.9-92.7]) | 240 (1.5%, [1.3-1.7]) | 1015 (6.2%, [5.8-6.6]) |
|  | Yorkshire and The Humber | 10547 (92.8%, [92.3-93.3]) | 155 (1.4%, [1.2-1.6]) | 661 (5.8%, [5.4-6.3]) |
| **IMD quintile** | 1 - most deprived | 14674 (86.7%, [86.2-87.2]) | 465 (2.7%, [2.5-3]) | 1785 (10.5%, [10.1-11]) |
|  | 2 | 24632 (89.7%, [89.3-90]) | 547 (2%, [1.8-2.2]) | 2283 (8.3%, [8-8.6]) |
|  | 3 | 34185 (91.9%, [91.6-92.2]) | 546 (1.5%, [1.4-1.6]) | 2468 (6.6%, [6.4-6.9]) |
|  | 4 | 39668 (93%, [92.8-93.3]) | 498 (1.2%, [1.1-1.3]) | 2473 (5.8%, [5.6-6]) |
|  | 5 - least deprived | 45156 (94.3%, [94.1-94.5]) | 420 (0.9%, [0.8-1]) | 2299 (4.8%, [4.6-5]) |
| **Key worker status** | Healthcare worker | 7663 (92.1%, [91.5-92.7]) | 226 (2.7%, [2.4-3.1]) | 432 (5.2%, [4.7-5.7]) |
|  | Care home worker | 1121 (88.5%, [86.7-90.2]) | 57 (4.5%, [3.5-5.8]) | 88 (7%, [5.7-8.5]) |
|  | Other key worker | 36727 (90%, [89.8-90.3]) | 776 (1.9%, [1.8-2]) | 3283 (8%, [7.8-8.3]) |
|  | Other worker (not key worker) | 53588 (91.1%, [90.9-91.3]) | 780 (1.3%, [1.2-1.4]) | 4465 (7.6%, [7.4-7.8]) |

**Figure 4.**
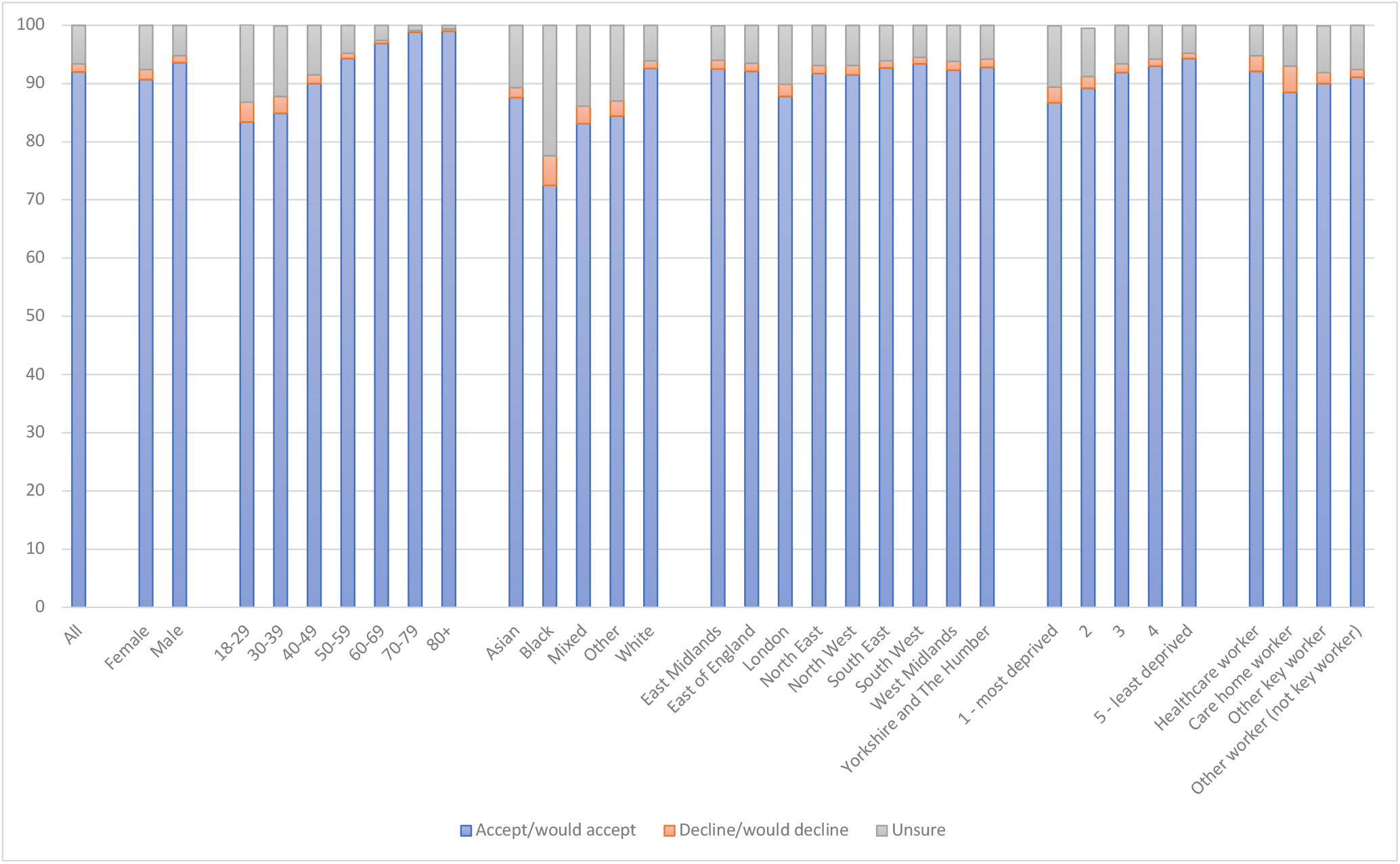
Vaccine confidence by key covariates.

## Discussion

In round 5 of the REACT-2 study during January and February 2021, we found increased population prevalence of anti-SARS-CoV-2 antibodies compared with November 2020, reflecting both high levels of infection during the second peak of the epidemic in England and response to the early stages of the national vaccination programme. In contrast to our previous surveys, the highest prevalence of antibodies was found in those aged 80 years and over, amongst whom over 90% had been vaccinated.

As previously reported with respect to the first wave, for unvaccinated individuals, we found increased prevalence among healthcare and care home workers, people of Black and Asian ethnicities, and those living in more deprived areas, as well as in London and among younger people (ages 18 to 29 years). (1) We also found that key workers such as those in education, public transport and other public-facing roles had higher antibody prevalence than non-key workers. These results indicate that variation and inequities in risk of infection noted in the first wave persisted into the second wave.

We show that confidence in the vaccine programme in England is generally very high, although lower in some groups including younger people and people of Black ethnicity. The slightly lower confidence in women may reflect the fact that vaccination is not currently advised in pregnancy,(6) and wider concerns about future fertility. To ensure that the vaccination programme is rolled out equitably to all sections of society, messaging of the benefits of the programme – to the individual, their family, contacts and wider society – needs to be made readily accessible to different communities.

The Phase 3 registration trial of the Pfizer-BioNTech vaccine demonstrated an efficacy of 95% against clinical disease seven days after the second vaccine dose.(19) The majority of participants in those studies were aged 16 to 55 years, with a smaller proportion of individuals aged over 65 years. In that study, clinical protection did not appear to differ materially in older age groups, but the small study numbers meant there was uncertainty around these estimates. Our results confirm a high prevalence of detectable antibody following two doses of Pfizer-BioNTech, consistent with the clinical protection seen in trials. High levels of antibody positivity were seen across all age groups after two doses, although lower in those 80 years and over than in those under 50 years. This may reflect higher rates of prior infection at younger ages or possibly reduced antibody response to vaccination in the oldest people consistent with some other vaccines.(20)

A single Pfizer-BioNTech vaccination was associated with high antibody positivity in those with previous suspected or confirmed COVID-19. It is recognised that, in sera from individuals infected with SARS-CoV-2 at least six months prior, neutralising antibody may be at a low level. However, when such individuals are given a single dose of an mRNA vaccination (either Pfizer-BioNTech/BNT162b2 or mRNA-1273) substantial, rapid increases in anti-Spike IgG are observed.(21–23) Given that these subsequent antibody titres may be at least as high as those in uninfected individuals given two vaccine doses, and the increased reactogenicity of second vaccine doses,(22) it has been suggested that those with established prior infection might be given lower priority access to vaccination (or longer delay in second doses of two dose regimens). Although the durability of primed responses to single dose in those previously infected is not yet known, France is among countries using prior infection to prioritise limited vaccine doses. The data here add support to that approach, particularly in younger age groups, though consideration needs to be given to the additional complexity to rapid vaccine roll out that this approach might require.

There are relatively limited data on the efficacy of either Pfizer-BioNTech or AstraZeneca/Oxford vaccines in older populations. At the time of our study, the number of individuals who had received the AstraZeneca/Oxford vaccine more than 21 days earlier was too low to allow meaningful analyses, although planned head-to-head trials will be able to address the comparative efficacy more clearly. A recent Israeli study found good responses to a single dose of Pfizer-BioNTech vaccination, with a suggestion that antibody titres were lower with increasing age.(24) A study of UK healthcare workers found an inverse correlation between age and anti-S antibody responses following a single dose and significantly higher anti-S responses in those aged under 50 compared to those over 50. (Prendecki 2021, in press) In those receiving a single dose of Pfizer-BioNTech vaccine, we observed decrease in antibody positivity with increasing age, although, as noted above, there were high levels of antibody positivity across all age groups after two doses.

There are a number of limitations to this study. While the use of self-administered lateral flow tests for population surveillance allows the rapid evaluation of large numbers of individuals in a cost-effective manner, these LFIA assays are generally less sensitive than laboratory assays.(12) In addition, they provide a threshold reading rather than a quantitative assessment of antibody response. As such, the estimates of antibody positivity here are likely to be lower than those obtained on laboratory platforms and it is unclear the extent to which antibody positivity, including from LFIA, correlates with protective immunity. However, we demonstrate that the detection of antibody on the test used correlates well with a threshold for neutralisation of live virus in in vitro assays. In addition, both Pfizer-BioNTech(25) and AstraZeneca/Oxford(26) vaccines generate antibody and T cell mediated immune responses such that vaccinated individuals may have T cell mediated protection even if antibody responses are not detected. LFIA tests may also be subject to errors when used at home, although we have found good usability in earlier work including in older people.(13) However, it is possible that poorer visual acuity in older people affects the ability to read the test result if there is a faint line.

It is a high priority to establish the relationship between antibody positivity and the subsequent risk of hospitalisation and/or death. Initial data from a cohort of UK healthcare workers suggests a single of Pfizer-BioNTech vaccination is associated with a 72% reduction in infection after 21 days.(9) In studies of individuals 80 years or over, a single dose of BNT162b2 is associated with a greater than 50% reduction in cases 28 days after vaccination, rising to 98% after second doses are given, emphasising the importance of second doses, particularly in older populations.(27)

The analysis here is limited to the of Pfizer-BioNTech vaccine and there were insufficient data for comparison with the AstraZeneca/Oxford vaccine. The data here suggest the optimum interval may need to be tailored to population groups, with a longer delay in second doses more appropriate for younger age groups and those with prior infection. In addition it is important to establish the relationship between antibody positivity following vaccination and the subsequent risk of hospitalisation and/or death in order to assess whether antibody response is a useful correlate of protection. Randomised trials to inform the optimum timing of first and second vaccinations are underway and, along with growing bodies of real-world evidence, will help inform challenging prioritisation decisions for national and international bodies.

## Data Availability

The data analysed or used, or both, in this study are not publicly available owing to governance restrictions.

## Data availability

Summary tabular data are provided with this paper.

## Acknowledgements

HW is a National Institute for Health Research (NIHR) Senior Investigator and acknowledges support from NIHR Biomedical Research Centre of Imperial College NHS Trust, NIHR School of Public Health Research, NIHR Applied Research Collaborative North West London, and Wellcome Trust (UNS32973). GC is supported by an NIHR Professorship. WSB is the Action Medical Research Professor, AD is an NIHR senior investigator and DA and PE are Emeritus NIHR Senior Investigators. SR acknowledges support from MRC Centre for Global Infectious Disease Analysis, National Institute for Health Research (NIHR) Health Protection Research Unit (HPRU), Wellcome Trust (200861/Z/16/Z, 200187/Z/15/Z), and Centres for Disease Control and Prevention (US, U01CK0005-01-02). PE is Director of the MRC Centre for Environment and Health (MR/L01341X/1, MR/S019669/1). PE acknowledges support from the NIHR Imperial Biomedical Research Centre and the NIHR HPRUs in Chemical and Radiation Threats and Hazards and in Environmental Exposures and Health, the British Heart Foundation Centre for Research Excellence at Imperial College London (RE/18/4/34215), Health Data Research UK (HDR UK) and the UK Dementia Research Institute at Imperial (MC_PC_17114). We thank The Huo Family Foundation for their support of our work on COVID-19. SD acknowledges support from NIHR Biomedical Research Centre of Imperial College NHS Trust.

We thank key collaborators on this work --Ipsos MORI: Stephen Finlay, John Kennedy, Kevin Pickering, Duncan Peskett, Sam Clemens and Kelly Beaver; Institute of Global Health Innovation at Imperial College London: Gianluca Fontana, Dr Hutan Ashrafian, Sutha Satkunarajah, Didi Thompson and Lenny Naar; the Imperial Patient Experience Research Centre and the REACT Public Advisory Panel; NHS Digital for access to the NHS Register.

## Supplementary material

**Table S1:** REACT-2 round 5, response rates.

|  | **Study 5 Round 5**<br>25 Jan - 08 Feb<br>2021 | % (of<br>sampled) | % (of<br>those<br>registered) |
| --- | --- | --- | --- |
| Sample invited | 600,018 |  |  |
| Registration - agreed to receive LFT<br>test | 194,762 | 32.5% |  |
| Symptom surveys done | 172,099 | 28.7% | 88.4% |
| Attempted LFT test | 159,983 | 26.7% | 82.1% |
| Completed LFT test | 157,698 | 26.3% | 81.0% |
| Valid LFT test result | 155,172 | 25.9% | 79.7% |

## Supplementary Methods

### Detailed Methods for healthcare worker study Study Participants

Eighty healthcare workers at Imperial College Healthcare NHS trust were recruited to the study between 23^rd^ December 2020 and 31^st^ January 2021, at the time of receiving their first dose of BNT162b2 vaccine. Seventy-two participants provided a subsequent blood sample at 21-28 days following vaccination and are included in this analysis. Data were collected on age and gender. Medical records of participants were not accessed. The study was approved by the Health Research Authority, Research Ethics Committee (Reference: 20/WA/0123).

### Serological testing

Serum was tested for antibodies to nucleocapsid protein (anti-NP) using the Abbott Architect SARS-CoV-2 IgG 2 step chemiluminescent immunoassay (CMIA) according to manufacturer’s instructions. This is a non-quantitative assay and samples were interpreted as positive or negative with a threshold index value of 1.4. Spike protein antibodies (anti-S) were detected using the Abbott Architect SARS-CoV-2 IgG Quant II CMIA. Anti-S antibody titres are quantitative with a threshold value for positivity of 50 AU/ml.

### In vitro live virus neutralisation assay

The ability of sera to neutralise SARS-CoV-2 virus was assessed by neutralisation assay on Vero cells. Sera were serially diluted in OptiPRO SFM (Life Technologies) and incubated for 1h at RT with 100 TCID_50_/well of SARS-CoV-2/England/IC19/2020 and transferred to 96-well plates pre-seeded with Vero-E6 cells. Serum dilutions were performed in duplicate. Plates were incubated at 37°C, 5% CO_2_ for 42 h before fixing cells in 4% PFA. Cells were treated with methanol 0.6% H_2_O_2_ and stained for 1h with a 1:3000 dilution of 40143-R019 rabbit mAb to SARS-CoV-2 nucleocapsid protein (Sino Biological). A 1:3000 dilution of sheep anti-rabbit HRP conjugate (Sigma) was then added for 1 h. TMB substrate (Europa Bioproducts) was added and developed for 20 mins before stopping the reaction with 1M HCl. Plates were read at 450nm and 620nm and the concentration of serum needed to reduce virus signal by 50% was calculated to give NT_50_ values.

### Statistical Analysis

Statistical analysis was conducted using Prism 9.0 (GraphPad Software Inc., San Diego, California). Unless otherwise stated, all data are reported as median with interquartile range. Where appropriate, Mann-Whitney U and Kruskal-Wallis tests were used to assess the difference between 2 or >2 groups, with Dunn’s post-hoc test to compare individual groups.

## Footnotes

1 https://www.imperial.ac.uk/medicine/research-and-impact/groups/react-study/

